# DNA capture and amplicon enrichment approaches for next-generation sequencing of *Mycoplasma genitalium* directly from clinical samples

**DOI:** 10.64898/2026.04.29.26351683

**Authors:** Jennifer Guiraud, Charlotte Héricé, Carla Balcon, Léo Gillet, Marie Gardette, Arabella Touati, Sabine Pereyre, Cécile Bébéar

## Abstract

Direct genome sequencing of *Mycoplasma genitalium* from clinical specimens is challenging due to the organism’s low bacterial load. We developed and compared two DNA enrichment strategies—amplicon-based and hybridisation capture-based—coupled with next-generation sequencing, and assessed the suitability of DNA capture for whole-genome sequencing and high-resolution molecular typing. The enrichment approaches, RNA bait hybridisation and targeted sequence amplification, were combined with paired-end sequencing on the Illumina iSeq 100. Method performance was evaluated in 89 *M. genitalium*-positive specimens across five genomic loci: 23S and 16S rRNA, and *parC* and *gyrA* for macrolide, tetracycline, and fluoroquinolone resistance, respectively, and *mgpB* for phylogenetic analysis.

Regarding antimicrobial resistance (AMR) determinants, the multiplex amplicon sequencing method demonstrated the highest sensitivity, at 92.1–98.9%. Although < 50% of samples were characterised using DNA capture, concordance between the two methods was excellent. Using DNA capture, 33.7% of specimens achieved a minimum coverage of 90% at 1× depth. Sequencing success was inversely correlated with human and bacterial DNA contamination, and the presence of low-quality reads. Multi-locus sequence typing schemes provided the highest discriminatory power while all three typing methods evaluated in this study, including whole-genome single-nucleotide polymorphism analysis, clustered together multidrug-resistant *M. genitalium* strains belonging to the *mgpB* genotype 159.

In conclusion, under the specific set of parameters evaluated in this study, targeted amplicon-based enrichment demonstrated superior accuracy and reliability for AMR detection and *mgpB* typing, whereas DNA capture offered a secondary approach for generating comprehensive genomic data from selected *M. genitalium*–positive specimens.

**Impact statement:** Culturing *Mycoplasma genitalium* from clinical specimens is challenging; consequently, epidemiological studies rely on molecular techniques. Whole-genome sequences have been obtained from a few *M. genitalium* clinical isolates, and direct sequencing from clinical specimens requires enrichment strategies to overcome the organism’s low bacterial load. Consequently, published data on antimicrobial resistance (AMR) mechanisms and phylogenetic relationships among circulating strains remain incomplete.

In our study, targeted amplicon-based enrichment provided high-resolution and highly sensitive AMR detection, enabling the identification of minority variants. In contrast, hybridisation capture–based enrichment showed lower sensitivity with a specific set of parameters, but permitted successful WGS in a subset of *M. genitalium*–positive specimens and supported the development of new typing schemes. Although amplicon-based enrichment remains a reliable approach for epidemiological studies, strategic application of DNA capture facilitates the generation of comprehensive genomic data even though performance is reduced in the context of low bacterial loads, high DNA contamination or suboptimal DNA quality.

## Introduction

*Mycoplasma genitalium* is a sexually transmitted pathogen that causes infections of the lower urogenital tract in men and women (1). Although *M. genitalium* is uncommon in the general population, its prevalence can reach up to 20% among men who have sex with men (MSM) who are living with human immune deficiency virus (HIV) or using HIV preexposure prophylaxis (2-4). The increasing prevalence of macrolide and fluoroquinolone resistance led to the recognition of *M. genitalium* as a major global health threat by the US Centers for Disease Control and Prevention in 2019 (5). More recently, the spread of dual-resistant *M. genitalium* clones has been reported in France, especially among MSM populations (6). Other therapeutic approaches are limited to the use of doxycycline, minocycline, or pristinamycin, which are associated with limited clinical success rates (7).

Due to its extremely slow growth rate in culture, which necessitates enriched or cell-based media, diagnosis and antimicrobial resistance (AMR) testing for *M. genitalium* are almost exclusively reliant on molecular techniques (1). Studies using phenotypic approaches, such as minimum inhibitory concentration determinations, remain limited, restricting the understanding of AMR mechanisms, particularly for drugs, such as pristinamycin and tetracyclines. Furthermore, epidemiological studies rely on culture-independent techniques. Current molecular typing methods for *M. genitalium* rely on polymorphism analysis within a single locus, namely the *mgpB* gene (MG191 locus), which encodes the major adhesin. Short tandem repeat analysis of the putative lipoprotein gene *MG309* has been used to investigate transmission dynamics within close sexual networks (8-10).

Whole-genome sequencing (WGS) of *M. genitalium*, performed only on a few clinical isolates to date, is increasingly recognised as critical for generating comprehensive data on the phylogenetic relationships between strains, identifying novel genetic markers, improving the understanding of transmission dynamics, and elucidating AMR mechanisms (11-13). However, a major limitation is the low bacterial load of *M. genitalium* in clinical specimens. To overcome these limitations, DNA enrichment techniques coupled with next-generation sequencing (NGS) have been developed. These methods have been applied to other sexually transmitted bacteria, notably *Chlamydia trachomatis*, *Treponema pallidum*, and *Neisseria gonorrhoeae* (14-21). Enrichment has enabled access to genomic data from clinical strains while circumventing the difficulties associated with culture.

Enrichment techniques are broadly based on polymerase chain reaction (PCR) amplicon-based enrichment or hybridisation capture–based enrichment. Using clinical samples, although the former approach is still largely limited to specific genomic regions, the latter can yield entire genome sequences, provided that sufficient DNA input is available. Targeted amplicon sequencing methods have been developed to detect macrolide and fluoroquinolone resistance in *M. genitalium* and to determine *mgpB* genotypes (22-24). However, the utility of DNA capture remains unclear, with only two previous studies reporting limited success and partial genome sequence recovery for *M. genitalium* (15, 25).

We assessed the performance of two distinct DNA enrichment techniques coupled with NGS for the detection of AMR determinants and the genotyping of *M. genitalium* directly from clinical specimens. In particular, the performance of amplicon enrichment and DNA capture approaches was compared across five genomic loci: the 23S rRNA gene associated with macrolide resistance, the *parC* and *gyrA* genes associated with fluoroquinolone resistance, the 16S rRNA gene associated with tetracycline resistance, and the *mgpB* gene used for phylogenetic analysis. Beyond targeted gene detection, we evaluated the suitability of DNA capture for WGS and for the development of high-resolution molecular typing schemes for *M. genitalium*.

## Methods

### Study design and participants

The French National Reference Centre for Bacterial Sexually Transmitted Infections (STI NRC) centralised molecular diagnostics for macrolide and fluoroquinolone resistance in *M. genitalium*–positive specimens obtained from patients in French microbiology laboratories. A questionnaire was used to collect demographic, clinical, and antibiotic treatment data from the participating laboratories.

We selected 89 remnant specimens collected as part of routine care and surveillance studies conducted by the STI NRC between 2021 and 2024, with a cycle threshold ≤ 30 using *M. genitalium* detection (Cobas CT/NG Test for Use on the Cobas 6800/8800 Systems) and macrolide resistance diagnostic kits (Macrolide-R/MG ELITe MGB Kit) (26). Genomic DNA was extracted from 500 µL of clinical samples using Genomic DNA and RNA Purification AXG20 columns and the NucleoBond Buffer Set III/IV (Macherey-Nagel Inc., Bethlehem, PA, USA). DNA extracts were quality controlled using a Qubit 4 fluorometer (Thermo Fisher Scientific, Waltham, MA, USA) to determine total DNA concentrations. *M. genitalium* bacterial load was quantified using an in-house quantitative TaqMan PCR targeting a 78-bp region of the MgPa adhesin gene (27). Two enrichment approaches—RNA bait hybridisation and targeted sequence amplification coupled with high-throughput sequencing—were used to investigate the genotype and antibiotic resistance profile of *M. genitalium* strains. DNA from the *M. genitalium* reference strain (ATCC 33530) was used as a positive control.

## Procedures

### WGS through DNA capture

Custom probes were designed by Agilent Technologies (Santa Clara, CA, USA) based on the reference genomes NC000908.2 (MG G37), CP003770.1 (M2321 isolate), CP003771.1 (M6282 isolate), CP003772.1 (M6320 isolate), and CP003773.1 (M2288 isolate), as well as raw sequences from genome sequencing of 31 *M. genitalium* isolates (BioProject PRJEB5172). The bait set comprised 11,762 120-mer RNA baits (Design ID S3347512). Sequence libraries were prepared using the Magnis NGS Prep System (Agilent Technologies) with the Magnis SureSelect XT HS Reagent Kit for Illumina paired-end platforms (Illumina, San Diego, CA, USA). For the initial step of enzymatic DNA fragmentation, an input volume of 14 µL was used because of the low DNA concentrations and the manufacturer’s recommendations. The final volume was adjusted accordingly to reach a total of 50 µL. The incubation step for enzymatic fragmentation was performed at 37°C for 15 min. During the second stage, library preparation, 12 PCR cycles were used for pre-hybridisation amplification. The probes were used at a 1:10 dilution. Hybridisation was performed overnight at 65°C. Post-hybridisation amplification was conducted using 22 PCR cycles. All libraries were quality controlled using a Qubit 4 fluorometer (Thermo Fisher Scientific) and a TapeStation system (Agilent technologies). Paired-end sequencing (2×150 bp) was performed on an Illumina iSeq100 platform (Illumina). To confirm the absence of contamination, *M. genitalium-*negative samples were included as negative controls and processed using the same protocol. Samples were multiplexed in six sequencing runs in batches of 8, 16, or up to 24 samples.

### Custom amplicon sequencing

The antibiotic resistance profile of *M. genitalium* strains was determined by amplification of molecular antibiotic targets, including regions of interest (ROI) within genes encoding 23S rRNA domain V for macrolide resistance, the quinolone resistance–determining regions of *parC* and *gyrA* for fluoroquinolone resistance, and helices 31 and 34 of the 16S rRNA gene for tetracycline resistance, using the primers listed in Supplementary table 1. All amplicons obtained from the same sample were pooled and then subjected to an Illumina tagmentation protocol (S beads) for library preparation (Illumina). Two independent sequencing runs of pooled amplicons were performed on an Illumina iSeq100 platform (Illumina). To confirm the absence of contamination, negative controls were processed using the same protocol. Samples with unsuccessful NGS and remaining DNA underwent Sanger sequencing. To determine the genotype of *M. genitalium* strains, the *mgpB* adhesin gene was amplified using the primer pair MgPa-1/MgPa-3 as previously described (8). Due to an incompatibility between the LightCycler master mix and the library preparation reagents, *mgpB* amplicons underwent Sanger sequencing.

### Bioinformatics

A custom Python pipeline was developed to process paired-end sequencing data generated using DNA capture and custom amplicon approaches for *M. genitalium*. The scripts are publicly available on GitHub (https://github.com/chah1337/DNA-Capture-VS-Amplicon-Mg.git). The pipeline was implemented in Python v3.12.9 using pysam v0.23.3, pandas v2.3.2, NumPy v2.3.3 and Biopython v1.85, under Red Hat Enterprise Linux 9.6. Raw reads were quality-filtered and trimmed using fastp v1.0.1 (minimum read length, 50 bp; minimum average quality, Q15), including adapter, poly-G and poly-X trimming and quality correction. For DNA capture data, 5-bp unique molecular identifiers (UMIs) were extracted from reads. Human-derived reads were removed using Bowtie2 v2.5.1. Remaining reads were mapped to the *M. genitalium* G37 reference genome (NC_000908.2) using BWA-MEM v0.7.19-r1273. BAM files were sorted, processed and indexed using samtools v1.23.1. Mapping statistics, mean sequencing depth and genome-wide and region-of-interest (ROI) coverage were calculated, including coverage at 1×, 5× and 10× depth. Variant calling was performed using FreeBayes v1.3.10 (ploidy = 2, minimum variant allele frequency = 5%, minimum alternate allele count = 2), followed by filtering and normalization using bcftools v1.23.1 and vt v0.57721. For AMR-associated loci (16S and 23S rRNA, *gyrA* and *parC*), consensus sequences were generated using samtools consensus, with a minimum depth threshold of 5×; positions with insufficient coverage were masked as N. To account for mixed populations, variants with allele frequencies ≥20% and <80% were additionally masked as N, whereas variants below 20% were disregarded and variants above 80% were retained as the majority allele. Variants were annotated using SnpEff v5.4c. AMR interpretation reliability was assessed using gene-specific coverage (>80%), mean sequencing depth (>10x), and minimum variant allele frequency (> 10%). A custom bioinformatics pipeline followed by manual inspection of BAM files in the Integrative Genomics Viewer (IGV) was used to check read alignments, depth, and potential artifacts (non-specific amplification). For *parC* and *gyrA* genes, only nonsynonymous mutations were reported.

For phylogenetic analyses, samples with whole-genome coverage >80% at 5× depth were retained; sample MG3 was excluded due to a suspected mixed population. SNPs were extracted from the complete genomic sequences for whole-genome SNP (wg-SNP) analysis using SNP-sites (28). Core-genome multilocus sequence typing (cg-MLST) was generated using chewBBACA (29). For the human *Mycoplasma*-related MLST scheme (hMy-MLST), consensus sequences were generated for 12 selected housekeeping genes (*gap, tufA, ftsY, ppa, gpmI, gyrB, gmk, glyA, atpA, glpK, adk* and *mgpB*) (30, 31). Consensus sequences for *mgpB* and MLST loci were extracted from DNA capture data using BED-defined genomic regions. Consensus sequences were generated with a minimum depth threshold of 5×, variants below 50% were disregarded, and majority allele variants were retained. For cg-MLST and hMy-MLST, consensus sequences of all target genes were concatenated and aligned using MAFFT (v7.526, --auto option) before phylogenetic tree inference. For *mgpB*, the full gene sequence was aligned individually. For wg-SNP, whole-genome SNP alignments were generated using snp-sites (v2.5.1). All alignments were used as input for RAxML-NG (v1.2.0) with the GTR+G model to infer maximum-likelihood phylogenetic trees. Phylogenetic trees were visualised and annotated using R (version 4.5.1) and the ggtree framework. Assignment of *mgpB* types was performed by comparing the *mgpB* consensus sequences with the *M. genitalium* PubMLST database (https://pubmlst.org/) using BLAST (32). Newly described *mgpB* types were submitted to PubMLST for curation and assignment of a genotype number.

### Comparative phylogenetic analysis

To assess the discriminatory power of each typing method, we calculated two indices: the Shannon diversity index and the Hunter–Gaston discriminatory index. To evaluate consistency and divergence among the four typing methods (*mgpB*, hMy-MLST, cg-MLST, and wg-SNP), a comparative phylogenetic tree analysis pipeline was implemented. For each phylogenetic tree, we calculated a patristic distance matrix, with the distance between two taxa defined as the sum of the branch lengths connecting them. The absolute mean difference between each pair of patristic distance matrices was then calculated. This generated a 4×4 global difference matrix, which was visualised as heatmaps and used to construct a hierarchical clustering dendrogram using average linkage. The values quantified differences between the distance matrices generated by the typing methods. In addition, to quantitatively assess the congruence and consistency between the different typing schemes, Robinson-Foulds (RF) and Kendall-Colijn (KC) distances were also calculated. These matrix distances were based on topological discrepancies between trees. Regarding absolute distance value differences, low values indicated closely related taxa with weak divergence along the trees and likely belonging to the same cluster whereas higher values reflected more divergent related taxa, likely belonging to different clusters depending on distinct typing method used.

### Statistical analysis

Fisher’s exact test and the chi-squared test were used for qualitative variables, whereas analysis of variance and Student’s t-test were applied to quantitative variables. Linear regression analysis of determinants associated with genome sequencing success was performed using RStudio (version 4.5.1; 2025-06-13 ucrt; RStudio, PBC, Boston, MA, USA). Multiplex amplicon sequencing was considered the reference for kappa coefficient determination. P-values < 0.05 were considered statistically significant. Data visualisation was performed using ggplot2 v3.4.4.

### Ethics statement

Remnants of specimens were preserved at the Centre de Ressource Biologique–Bordeaux Biothèque Santé of Bordeaux University Hospital under collection number BB-0033-00094 and authorisation AC-2014-2166 from the French Ministry of Higher Education and Research. No information regarding patient identity was included, and all patient data were reported anonymously. In accordance with Article 6.1 of the General Data Protection Regulation (GDPR), which took effect on May 25, 2018, the data processing carried out by the National Reference Centres (NRCs) in the course of their duties is based on the performance of a task carried out in the public interest (Art. L 1413-3, R. 1413.46 et seq. of the Public Health Code (CSP)). This legal basis exempts the NRCs from the requirement to obtain prior consent from patients.

## Results

In total, 89 *M. genitalium*–positive and 4 *M. genitalium*–negative samples were subjected to both hybridisation capture–based and PCR amplicon–based enrichment, followed by Illumina sequencing. The positive samples were collected from 30 women and 59 men between 2021 and 2024 and comprised 58 urine samples (65.2%), 26 cervicovaginal swabs (29.2%), and 5 rectal swabs (5.6%) (Supplementary table 2). The median bacterial load obtained using in-house quantitative PCR was 2,300 genome copies/µL (interquartile range [IQR] 600–17,300), corresponding to a median cycle threshold (Ct) value of 30.65 (IQR 27.8–32.6) and a median DNA concentration of 9.1 ng/µL (IQR 4.2–19.2) (Supplementary table 2). At baseline, 23S rRNA sequence data were available for these samples, revealing a macrolide resistance prevalence of 58.4% (52/89) (Supplementary table 3). All sample metadata and metrics are available in the Supplementary table 2.

Total read counts were significantly higher with DNA capture than with amplicon sequencing (Supplementary figure 1). However, amplicon sequencing yielded to higher number of reads mapped specifically to AMR-determining regions.

Regarding AMR determinants, multiplex amplicon sequencing demonstrated high sensitivity for target gene detection, ranging from 92.1% (82/89) for the 16S rRNA gene to 98.9% (88/89) for the 23S rRNA, *parC*, and *gyrA* genes (Table 1). Amplicon derived results showed macrolide resistance–associated mutations in 60.2% (53/88) of samples, with the most frequent mutation being A2058T (29/53, 54.7%), followed by A2059G (n = 22) and A2058G (n = 6) (*Escherichia coli* numbering, Supplementary table 3). Of the 53 samples harbouring 23S rRNA mutations, 8 (15.1%) contained mixed sequences; in 4 of these, mutant and wild-type variants co-existed. Commercial macrolide resistance diagnostic kits failed to detect the A2059G variant, which was present at a 50% allelic frequency in sample 53 (Supplementary table 3). Fluoroquinolone resistance–associated mutations in ParC were observed in 36.4% (32/88) of samples, predominantly S83I (27/32, 84.4%), followed by D87N (n = 3), S83R (n = 1), and D87Y (n = 1) (*M. genitalium* numbering). Of the 32 samples with ParC mutations, 3 (9.4%) harboured mixed sequences of S83I and wild-type alleles, and 4 (12.5%) contained an additional GyrA mutation (M95I or D99G/N), of which 3 had mixed sequences combining mutated and wild-type GyrA alleles (75%). Among the three samples with 16S rRNA single-nucleotide polymorphisms (3/82, 3.6%), one *M. genitalium* strain carried the combination of G966T and G967T (*E. coli* numbering), whereas two contained mixed sequences consisting of 50% wild-type and 50% mutated alleles with both adjacent mutations, G966T and G967T (Supplementary table 3).

**Table 1.** Sensitivity of two enrichment and sequencing approaches for detection of antibiotic resistance target genes directly in clinical samples.

| **Antibiotics** | **Targeted genes** | **No. of samples tested** | **Amplicon sequencing** |  | **DNA capture** |  | ***p*-value<sup>a</sup>** |
|---|---|---|---|---|---|---|---|
|  |  |  | **Amplified n** | **Amplified % [IC95]** | **Amplified n** | **Amplified % [IC95]** |  |
| Macrolides | 23S rRNA | 89 | 88 | 98.9 [93.9-99.8] | 37 | 41.6 [31.9-52.0] | <0.001 |
| Fluoroquinolones | *parC* | 89 | 88 | 98.9 [93.9-99.8] | 37 | 41.6 [31.9-52.0] | <0.001 |
|  | *gyrA* | 89 | 88 | 98.9 [93.9-99.8] | 41 | 46.1 [36.1-56.4] | <0.001 |
| Tetracyclines | 16S rRNA | 89 | 82 | 92.1 [84.6-96.1] | 35 | 39.3 [29.8-49.7] | <0.001 |

<sup>a</sup>Chi-square test.

The sensitivity of DNA capture for detecting AMR was significantly lower than that of the multiplex amplicon sequencing method (P < 0.001; table 1). Overall, DNA capture demonstrated low and variable sensitivity across the target genes, ranging from 39.3% (35/89) for the 16S rRNA gene to 41.6% (37/89) for the 23S rRNA and *parC* genes, and 46.1% (41/89) for the *gyrA* gene. However, when DNA capture yielded a result, perfect concordance between the two methods was observed for the detection of resistance-associated mutations across all four targets (κ = 1; table 2).

**Table 2.** Clinical performances of the two enrichment and sequencing approaches in samples amplified with both methods.

| **Antibiotics** | **Targeted genes** | **No. of amplified samples tested** | **Identified as resistant by amplicon sequencing method** |  | **Identified as susceptible by amplicon sequencing method** |  | **κ<sup>a</sup>** |
|---|---|---|---|---|---|---|---|
|  |  |  | **Results obtained using DNA capture** |  |  |  |  |
|  |  |  | **Resistant** | **Susceptible** | **Resistant** | **Susceptible** |  |
| Macrolides | 23S rRNA | 37 | 26 | 0 | 0 | 11 | 1 |
| Fluoroquinolones | *parC* | 37 | 15 | 0 | 0 | 22 | 1 |
|  | *gyrA* | 41 | 2 | 0 | 0 | 39 | 1 |
| Tetracyclines | 16S rRNA | 34 | 2 | 0 | 0 | 32 | 1 |

<sup>a</sup>K, kappa concordance test.

The *mpg*B-based typing method identified genotypes in 77 (86.5%) *M. genitalium*–positive specimens using the amplicon sequencing method and in 52 (58.4%) using DNA capture. Sample 3 harboured a mixed *mgpB* sequence (Supplementary table 3). All *mgpB* types identified by the two methods were fully concordant. When the results were combined, in total 26 distinct *mgpB* types were identified, including 7 newly described types. Of the selected samples, types 159 (21/79, 26.6%), 4 (13/79, 16.4%), and 7 (12/79, 15.2%) were the most prevalent.

Using the DNA capture enrichment and Illumina sequencing approach, 30 specimens (33.7%) achieved a minimum breadth of coverage of ≥ 90% at 1× depth, 12 (13.5%) achieved ≥ 90% coverage at 5× depth, and 7 (7.8%) achieved > 90% coverage at > 10× depth (Supplementary table 2).

Although a positive association was observed between *M. genitalium* load and breadth of coverage, the observed variation in coverage is likely driven by other factors (Supplementary figure 2). To understand factors influencing WGS success better, we compared variables between samples grouped by breadth of coverage below or above 80% at 5× depth. A total of 23 specimens met this criterion and constituted the group with “successful WGS” results. The median *M. genitalium* inoculum and total DNA input concentration did not differ significantly between samples with successful and unsuccessful WGS (P > 0.05) (Supplementary figure 3). However, the median number of clean reads was significantly high in samples with successful WGS (418,960 vs 74,190; p < 0.0001). Notably, WGS success was associated with a significantly high percentage of on-target reads (reads mapped to the *M. genitalium* G37 reference genome) (61.8% vs 20.9%; P < 0.0001; figure 1). The percentages of short and low-quality reads were significantly lower in successful WGS samples compared to unsuccessful WGS samples (1.8% vs 5.6% and 1.0% vs 2.2%, respectively; P < 0.0001 and 0.01). Although the median percentage of unmapped reads—corresponding to DNA from other bacteria—was significantly lower in successful WGS samples compared to unsuccessful WGS samples (2.3% vs 3.6%; P < 0.01), the median percentage of human host–mapped reads was similar in both groups (21.7% vs 21.1%) (Figure 1). Overall, approximately 93,034 on-target reads were required to achieve ≥ 80% breadth of coverage at 5× depth (Figure 2).

**Figure 1.**
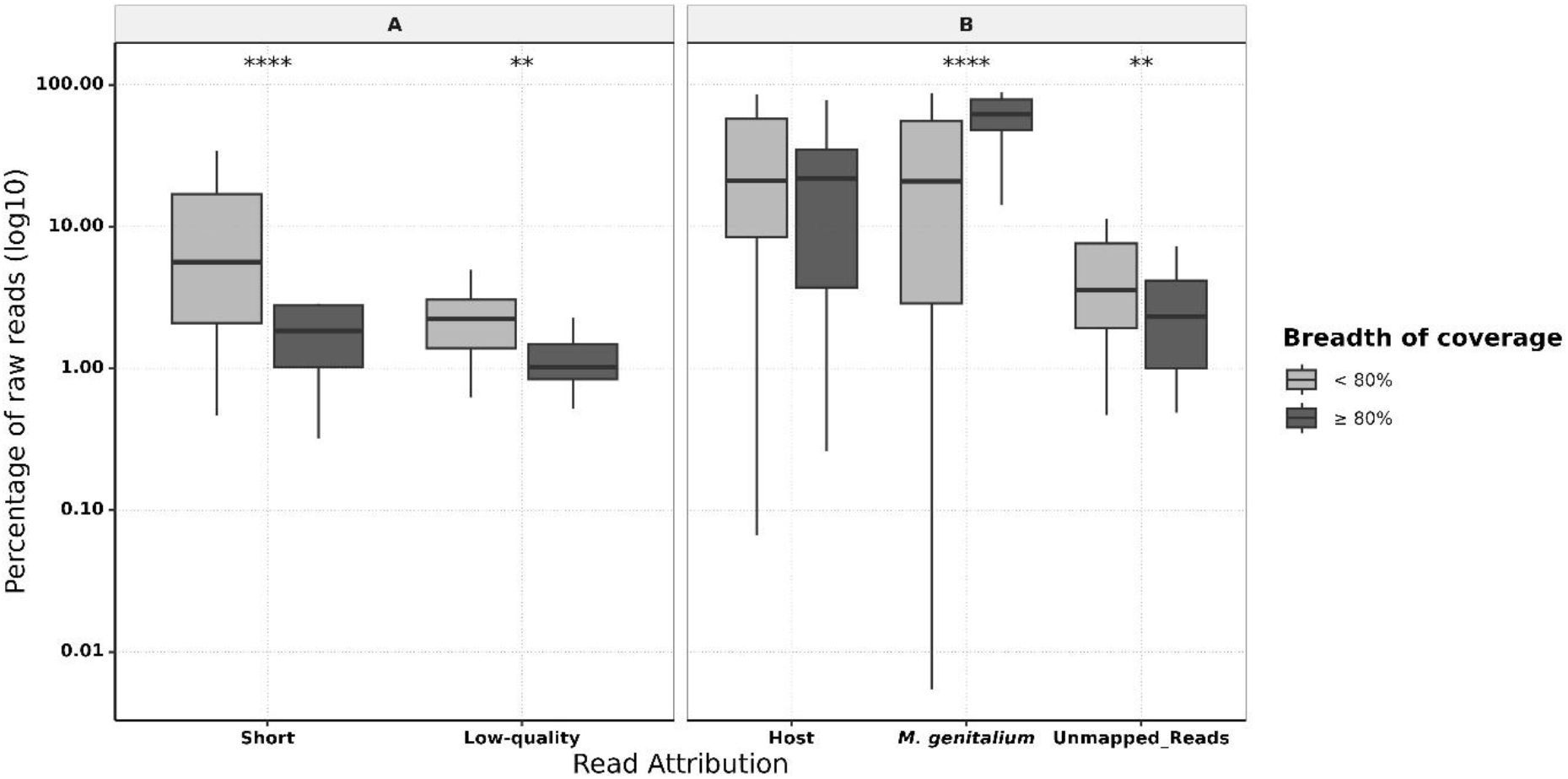
Impact of read quality and mapping distribution on *M. genitalium* whole-genome sequencing coverage. The box plots illustrate the distribution of read percentages categorized by their quality and attribution (host mapped reads, reads mapped on *M. genitalium* genome, unmapped reads). The data are stratified according to the breadth of coverage achieved at 5× (light grey: < 80%; dark grey: ≥ 80%). (A) The percentage of short and low-quality reads was calculated using the total raw read count. (B) The percentage of on- (*M. genitalium*) and off-target (host and unmapped) reads was calculated using the total clean read count. The horizontal lines within each box represent the median, while the whiskers indicate the range. Statistical significance markers are defined as: ∗∗p<0.01, and ∗∗∗∗p<0.0001. The absence of asterisks indicated that no significant differences in distribution were observed between the groups.

**Figure 2.**
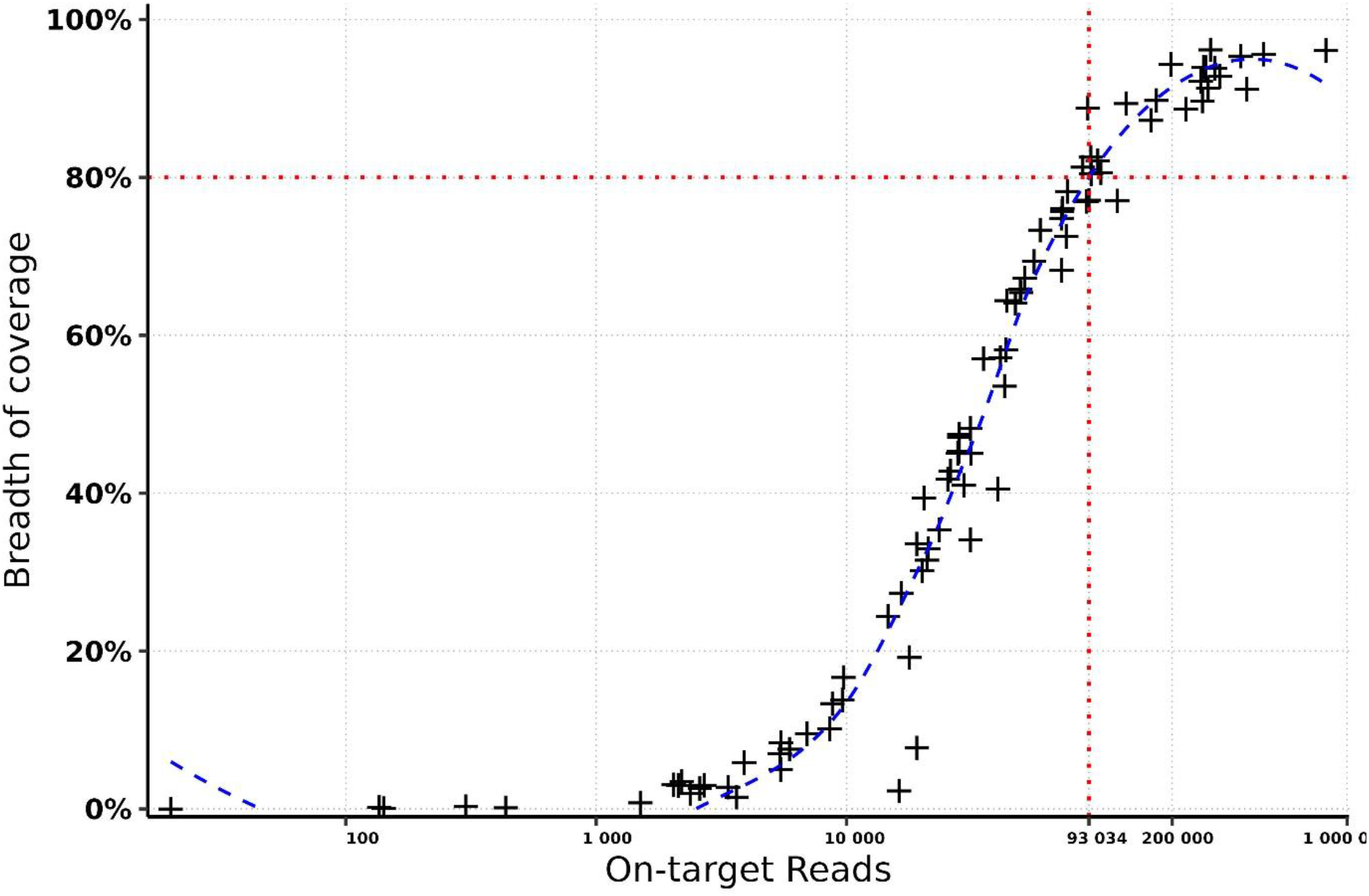
Breadth of coverage (5× depth) as a function of on-target read count. The trendline indicated that 93,034 on-target reads were necessary to achieve 80% breadth of coverage at 5× depth.

The median percentage genomic reference sites with at least 5 read coverage was significantly lower in vaginal swabs (15.2%) compared to urine samples (61.3%; P < 0.01; Figure 3). The difference was not significant when compared to anal swabs, likely because of the limited sample size (n = 5). This reduction in coverage correlated with a significantly higher proportion of host-mapped reads in vaginal swabs (38.5%) compared to urine samples (15.4%; P < 0.01) or anal swabs (5.1%; P < 0.05) (Figure 3). Although the percentage of unmapped reads remained relatively stable across all samples, at 2.3-4.9%, a lower median percentage of on-target reads was observed in vaginal swabs (7.6%) compared to urine samples (49.8%, P < 0.01) and anal swabs (73.3%; P < 0.05). These variations in coverage were independent of read count, bacterial inoculum, and total DNA concentration, as all quantitative metrics were comparable across the different sample types (Supplementary figure 4). Overall, these findings demonstrate a direct association between WGS success after DNA capture enrichment and the proportion of on-target reads, together with an inverse association with human and bacterial DNA contamination, and low-quality sequences.

**Figure 3.**
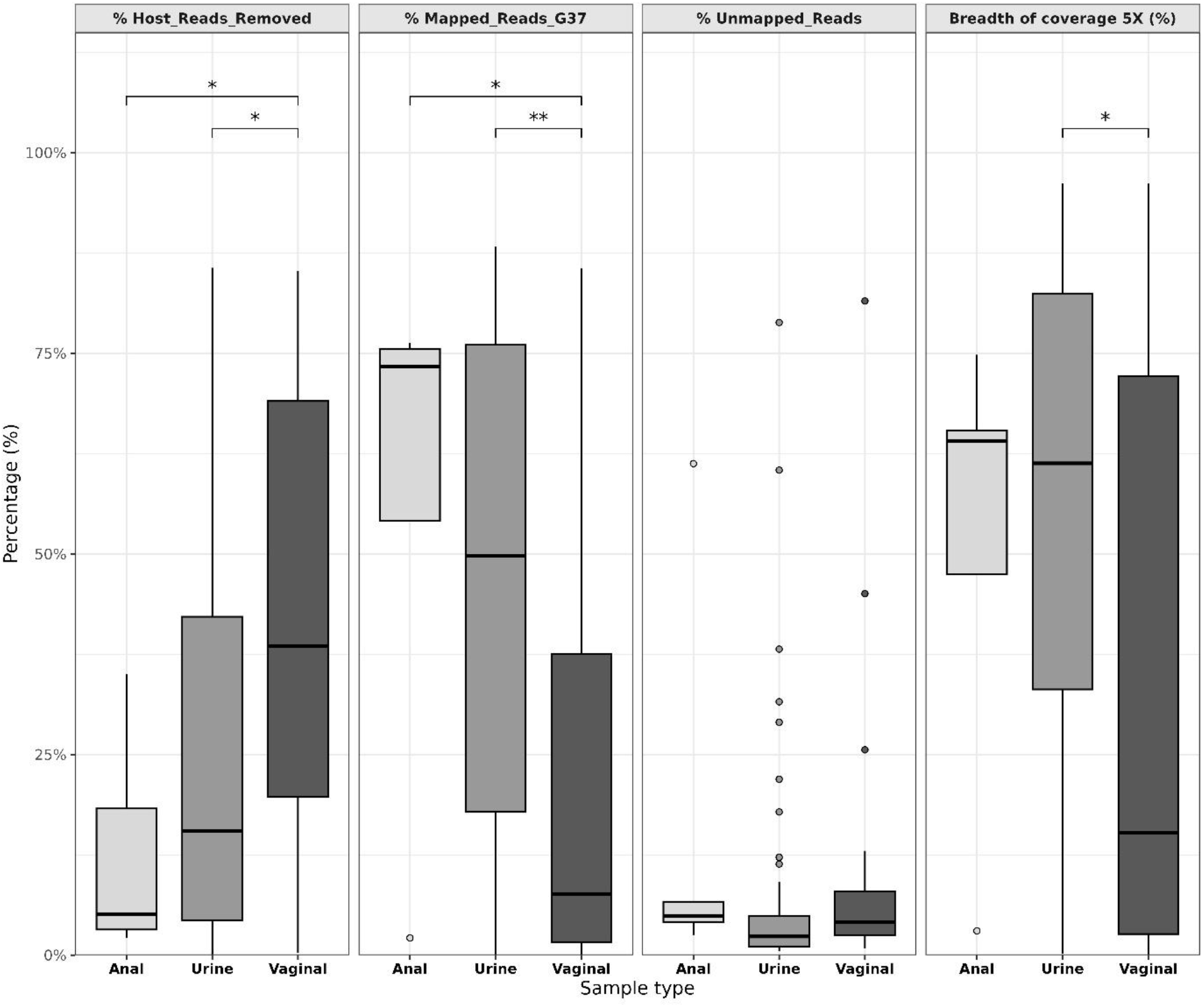
Comparison of sequencing metrics across clinical sample types. The box plots illustrate the distribution of sequencing data for anal (n=5), urine (n=58), and vaginal (n=26) samples across four key metrics: percentage of host mapped reads, percentage of reference mapped reads, percentage of unmapped reads and breadth of coverage at 5× depth. The horizontal lines within each box represent the median, while the whiskers indicate the range. Statistical significance markers are defined as: ∗p<0.05, ∗∗p<0.01. The absence of asterisks indicated that no significant differences in distribution were observed between the groups.

Of the 23 samples with a minimum breadth of coverage of 80% at 5× depth, 22 were retained for further phylogenetic analysis. Sample 3 was excluded because of the presence of mixed populations. The *mgpB* typing results were compared with those obtained using three molecular typing approaches: wg-SNP, cg-MLST, and hMy-MLST (Figure 4).

**Figure 4.**
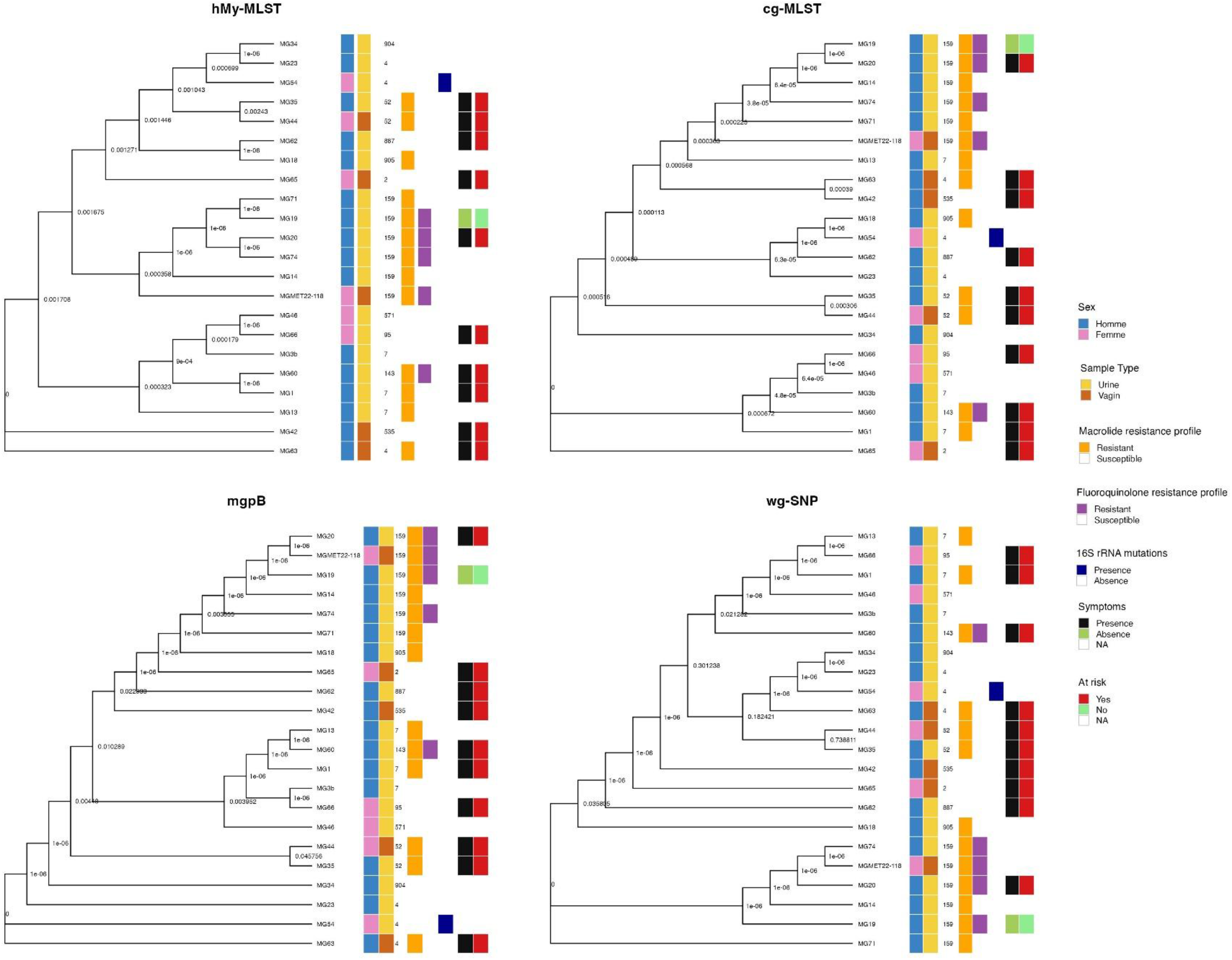
Maximum likelihood phylogenetic analysis of 22 *M. genitalium* strains based on the four typing methods (hMy-MLST, cg-MLST, *mgpB*, and wg-SNP). These trees represented the genotype distribution of 22 *M. genitalium*–positive specimens. The trees were inferred using the maximum-likelihood method based on either human *Mycoplasma* multi-locus sequence typing (hMy-MLST) profile, core-genome multi-locus sequence typing profile (cg-MLST), *mgpB* gene polymorphisms, and whole genome single nucleotide polymorphisms analysis (wg-SNP). The phylogenetic trees were annotated with the specimen name, patient sex, sample type, *mgpB* type, macrolide and fluoroquinolone susceptibility profiles, presence/absence of 16S rRNA mutations, presence/absence of symptoms and occurrence of at-risk sexual behaviour (person living with HIV or users for HIV pre-exposure prophylaxis).

The hMy-MLST scheme included 12 housekeeping genes previously used for typing *Mycoplasma hominis* and *Mycoplasma pneumoniae* strains (30, 31). Conversely, the *in silico*– constructed cg-MLST scheme developed in our study was based on 43 selected coding DNA sequences (CDSs) (Supplementary table 4). The resolving power of the four methods was further compared using discriminatory indices. The highest resolution was obtained using cg-MLST (Shannon index: 3.09, Hunter–Gaston index: 1.0), followed by hMy-MLST (2.72, 0.96), and the wg-SNP and *mgpB* typing methods (both 2.23, 0.90). Consistency and divergence were evaluated by three tree comparing metrics: patristic, Robinson-Foulds (RF), and Kendall-Colijn (KC) distances. Absolute mean differences between each pair of distance were visualised as a hierarchical clustering dendrogram and/or heatmap (Figure 5, supplementary figures 5, and 6).

**Figure 5.**
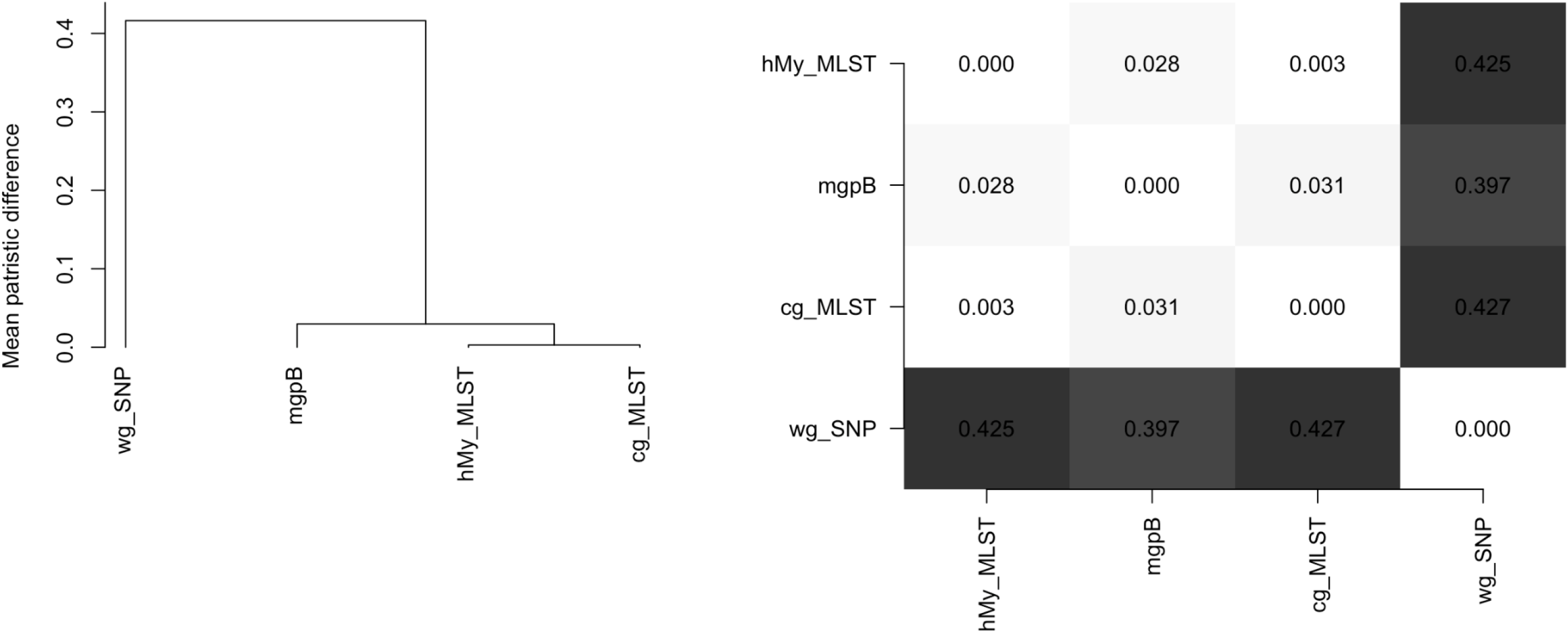
Comparison of phylogenetic tree topologies across four typing methods (*mgpB*, wg-SNP, hMy-MLST, cg-MLST) using patristic distances. The dendrogram (left) represents the hierarchical clustering of the four phylogenetic methods (hMy-MLST, *mgpB*, cg-MLST, and wg-SNP) based on the mean patristic distance (See materials and methods for details) between their respective matrices. Methods clustering closer together exhibit greater similarity in their phylogenetic reconstructions. The heatmap (right) visualizes the mean patristic distance between each pair of methods, with a color gradient ranging from white (lower distance, higher similarity) to dark grey (higher distance, greater divergence). This combined visualization provides an integrated overview of the relative similarity and divergence among the four methods.

Across all tree comparison metrics, lower distance values were observed between the two MLST approaches, indicating higher topological congruence. Conversely, the highest patristic distance values were obtained when comparing wg-SNP analysis to the remaining methods, underscoring its greater divergence (Figure 5). The *mgpB*-based typing method did not show exact concordance with either MLST scheme. Notably, several *M. genitalium* strains within the same *mgpB* type (types 4 and 7) did not cluster together when using the MLST approaches (Figure 4). However, all six *M. genitalium* strains belonging to *mgpB* type 159 clustered together across all three typing methods evaluated in this study. The KC and RF distance metrics, which account for differences in tree topology, exhibited low distance values between wg-SNP and *mgpB*-based phylogenies (Supplementary figures 5 and 6). Overall, no significant association was observed between genotypes and patient demographics, clinical infection outcomes, or the AMR profiles of the strains (Figure 4).

## Discussion

We evaluated the performance of two enrichment approaches for NGS of 89 *M. genitalium*– positive clinical specimens. Although NGS-based assays using amplicon enrichment for the 23S rRNA, *parC*, *gyrA*, and *mgpB* genes have been previously reported, we additionally screened for 16S rRNA mutations (22-24). Our study represents only the third to focus on WGS of *M. genitalium* directly from clinical specimens (15, 25).

Despite the greater total sequencing effort allocated to whole genome sequencing, a comparison of data from both enrichment approaches demonstrated significantly higher ROI-mapped read counts and greater sensitivity for multiplex amplicon sequencing in detecting AMR determinants compared to DNA capture. Despite this difference, concordance between the two methods was excellent. Our findings are consistent with those of Plummer et al. (23), who achieved success rates of 75–96% in 52 *M. genitalium*–positive specimens using a custom amplicon sequencing approach targeting the 23S rRNA, *mgpB, parC,* and *gyrA* genes. In contrast, Chiribau et al. (22) reported a lower success rate (68.5%; 98/143) with an alternative amplicon method targeting seven AMR-associated loci (23S rRNA, *rplD, rplV, parC, parE, gyrA,* and *gyrB*). These findings confirm that amplicon-based enrichment for NGS is an accurate and reliable method for epidemiological studies focusing on AMR testing (22-24). In addition, a PCR-based tiling approach using the Nanopore MinION platform has recently been developed for *M. genitalium* WGS (13). Although their method currently relies on cultured isolates rather than direct clinical samples, coupling amplicon-based enrichment with long-read sequencing represents a promising strategy for accessing genomic data for *M. genitalium* directly from clinical specimens. Indeed, a similar strategy has already been successfully validated using *Mycobacterium tuberculosis*-positive specimens (33). Notably, amplicon-based enrichment facilitates generation of more comprehensive AMR data, including the identification of minority variants, as evidenced by the occurrence of mixed sequences (23). Moreover, access to genomic data may provide insights into AMR mechanisms, such as acquired resistance to pristinamycin and tetracyclines (34-36). A key question remains regarding whether minority resistance variants or previously uncharacterised single-nucleotide polymorphisms contribute to clinical therapeutic failure. The fastidious growth requirements of *M. genitalium* continue to pose challenges for studying its AMR.

In our study, although one-third of specimens achieved a minimum coverage of 90% at 1× depth using the DNA capture approach, high-depth complete genomes were not recovered. These findings are consistent with those reported by Buttner et al. (15), who did not obtain any complete genome at > 10× depth. In contrast, Azzato et al. achieved a higher genome recovery rate, obtaining ≥ 90% coverage at 5× depth for 50.8% (189/372) of *M. genitalium*–positive specimens (25), compared with 13.5% (12/89) in the present study. Our findings indicate that sequencing performance was not solely correlated with *M. genitalium* inoculum in clinical specimens. However, strong associations between bacterial load and WGS success have been reported for other pathogens using DNA capture (14, 15, 17, 37). For instance, Clark et al. (37) demonstrated that clinical samples with Ct values ≤ 26 were likely to yield genomes of acceptable quality for *Neisseria meningitidis*. Similarly, Golparian et al. (18) reported a WGS success rate of 97.3% in selected *N. gonorrhoeae*–positive specimens characterised by a high inoculum (Ct < 20). In contrast, such high bacterial loads are extremely rare for *M. genitalium* in clinical practice.

We observed that the proportion of host-mapped reads negatively affected WGS coverage, whereas the proportion of commensal or contaminating bacterial DNA may also have had an impact on sequencing success. Vaginal swabs generally yielded suboptimal coverage due to significant human genomic DNA contamination, whereas urine samples were the most robust and reliable specimen type for WGS of *M. genitalium* using DNA capture. These findings are consistent with previous studies indicating that a higher bacterial load is required from vaginal swabs than from urine samples to obtain a complete genome sequence via DNA capture (15, 17). Considering the observed inverse relationship between host DNA content and WGS coverage, the use of host DNA depletion kits may be advisable (15). This approach could increase the relative proportion of *M. genitalium* DNA and improve WGS success, particularly when processing challenging specimens such as vaginal swabs.

Furthermore, we observed that the proportion of short- and low-quality reads was higher in samples with unsuccessful WGS compared with the proportion in samples with successful WGS. Moreover, the total read count was significantly lower in this group, indicating that samples with unsuccessful WGS contained a lower overall quantity and quality of *M. genitalium* DNA compared to those with successful WGS. Moreover, the exclusive use of clinical specimens collected as part of routine care and stored in lysis buffer may have introduced additional bias in our dataset. The use of native clinical specimens is recommended to minimise DNA fragmentation and optimise WGS success. Because DNA extraction methodology may influence the yields of short- and low-quality DNA, a comparative evaluation of different extraction methods is needed. Although the Magnis NGS Prep system reduces hands-on time and turnaround time, improving method performance, further technical optimisation is required and should be evaluated on *M. genitalium*–positive specimens (18). Potential improvements include designing a greater number of RNA probes, increasing the representation of probes targeting AMR loci, GC-content boosting, and testing lower hybridisation temperatures (15, 18). Refining the hybridisation capture to focus solely on AMR-related genes and specific typing loci may enhance detection sensitivity. In addition, high-throughput platforms could be utilized to maximize read depth and optimize multiplexing density. Overall, the sensitivity of the hybridization capture–based enrichment approach is highly dependent on sample quality and input characteristics. This performance variability may be influenced by several compounding factors, including *M. genitalium* bacterial load, sample storage conditions, DNA extraction quality, the burden of off-target host DNA, the choice of sequencing platform, and the integration of the automated Magnis NGS Prep System. Despite these operational constraints, the DNA capture enrichment approach has facilitated *M. genitalium* WGS directly from clinical specimens, enabling the development of novel typing schemes and the identification of potential epidemiological markers. Although MLST schemes based on 12 or 43 CDSs produced closely clustered phylogenies, molecular typing based on *mgpB* polymorphism analysis resulted in distinct patterns, indicating that single-gene typing may mask underlying genomic links. These findings are consistent with those of Azzato et al. (25), who reported poor concordance between *mgpB* typing and broader genomic population structures. Specific genetic variability in the MgPa operon can confound true evolutionary relationships in *M. genitalium* typing, implying that genome-based schemes may provide a more robust alternative. However, all three typing methods evaluated in this study confirmed the close genetic relatedness of *M. genitalium* strains belonging to *mgpB* type 159—a genotype associated with the dissemination of dual resistance to macrolides and fluoroquinolones in the MSM population (6). Despite potential sample selection bias, *mgpB* type 159 remained highly prevalent among the analysed specimens, emphasising a substantial multidrug-resistant threat in *M. genitalium*. While *mgpB* and wg-SNP analyses yielded distinct clustering patterns compared with the groupings of hMy-MLST and cg-MLST, this may primarily reflect differences in the genomic data fraction interrogated. However, we cannot exclude that recombination events can also potentially introduce confounding factors into evolutionary and phylogenetic inferences. The high topological consistency and discriminatory power suggest that the MLST schemes remain suitable for broader epidemiological surveillance. Ultimately, the number of CDSs included in our cg-MLST scheme could be expanded by utilizing a dataset with higher genome coverage. Further validation of the proposed typing schemes will require analyses of larger sample sets and a reassessment of minimum coverage thresholds.

Our study has limitations. First, only five anal swabs were analysed, limiting the interpretability of findings from this type of specimen. Second, because our RNA probe design was based on a limited number of *M. genitalium* genome sequences, the enrichment step may have introduced potential biases in DNA capture. Third, because this work focused specifically on target-enrichment efficiency, baseline metagenomic sequencing was not performed; hence, the initial ratio of host to non-host DNA in the raw samples was not established.

In conclusion, our findings emphasise the critical role of enrichment strategies for NGS of *M. genitalium* directly from clinical specimens. Under the specific set of parameters evaluated in this study, amplicon-based enrichment demonstrated superior accuracy and reliability for epidemiological investigations focused on AMR and *mgpB* typing, whereas DNA capture offered a secondary approach for generating comprehensive genomic data from selected specimens, particularly those with high *M. genitalium* load and optimal DNA quality. Notably, such genomic data are essential for identifying newly emerging and resistant clones within sexual networks, as exemplified by the dual-resistant *mgpB* type 159, whose clonality was confirmed in our study.

## Supporting information

Supplementary figures from 1 to 6

Supplementary tables from 1 to 5

## Data Availability

All Illumina raw sequence data is deposited with the European Nucleotide Archive under study number PRJEB111308. Newly characterized mgpB types were registered in the PubMLST database (IDs 902-908).

https://github.com/chah1337/DNA-Capture-VS-Amplicon-Mg.git

## Titles of supplementary tables and figures

**Supplementary figure 1. Comparison of read count produced with the two enrichment approaches (amplicon sequencing and DNA capture).**

**Supplementary figure 2. Relationship between *M. genitalium* load and genome coverage**

**Supplementary figure 3. Impact of clean read count, total DNA concentration, and *M. genitalium* inoculum on *M. genitalium* whole-genome sequencing coverage.**

**Supplementary figure 4. Clean read count, total DNA concentration, and *M. genitalium* inoculum distribution in specimens across different specimen types.**

**Supplementary figure 5. Comparison of phylogenetic tree topologies across four typing methods using Robinson-Foulds distances.**

**Supplementary figure 6. Comparison of phylogenetic tree topologies across four typing methods using Kendall-Colijn distances.**

**Supplementary table 1. Primers used for amplification of antibiotic resistance targets and *mgpB* gene.**

**Supplementary table 2. Sample metadata, accession numbers and sequencing metrics.**

**Supplementary table 3. Concordance of antimicrobial resistance determinant detection between amplicon sequencing and DNA capture methods.**

**Supplementary table 4. List of coding DNA sequences included in the core genome multi-locus sequences typing scheme (cg-MLST) and human *Mycoplasma* (hMy-MLST) multi-locus sequences typing schemes.**

## Data Summary

All Illumina raw sequence data is deposited with the European Nucleotide Archive under study number PRJEB111308. Newly characterized *mgpB* types were registered in the PubMLST database (IDs 902–908). The scripts used to bioinformatic, phylogenetic and statistical analysis are publicly available on GitHub (https://github.com/chah1337/DNA-Capture-VS-Amplicon-Mg.git).

## Authors contributions

Cécile Bébéar, Jennifer Guiraud, and Sabine Pereyre conceptualised the study. Jennifer Guiraud, Carla Balcon, Charlotte Héricé, Léo Gillet, Marie Gardette and Arabella Touati were responsible for data curation, which involved direct assessment and verification of the data. Cécile Bébéar and Jennifer Guiraud made the decision to submit the manuscript and wrote the original draft. Charlotte Héricé, Léo Gillet, and Jennifer Guiraud performed the bioinformatic and statistical analysis. All co-authors edited the manuscript and approved the final version.

## Acknowledgements

We gratefully acknowledge Chloé Le Roy for her technical assistance on this project.

## Editing

The English in this document has been checked by at least two professional editors, both native speakers of English. For a certificate, please see: http://www.textcheck.com/certificate/bYN64w

