## Supplementary figures from 1 to 6 for "DNA capture and amplicon enrichment approaches for next-generation sequencing of *Mycoplasma genitalium* directly from clinical samples"

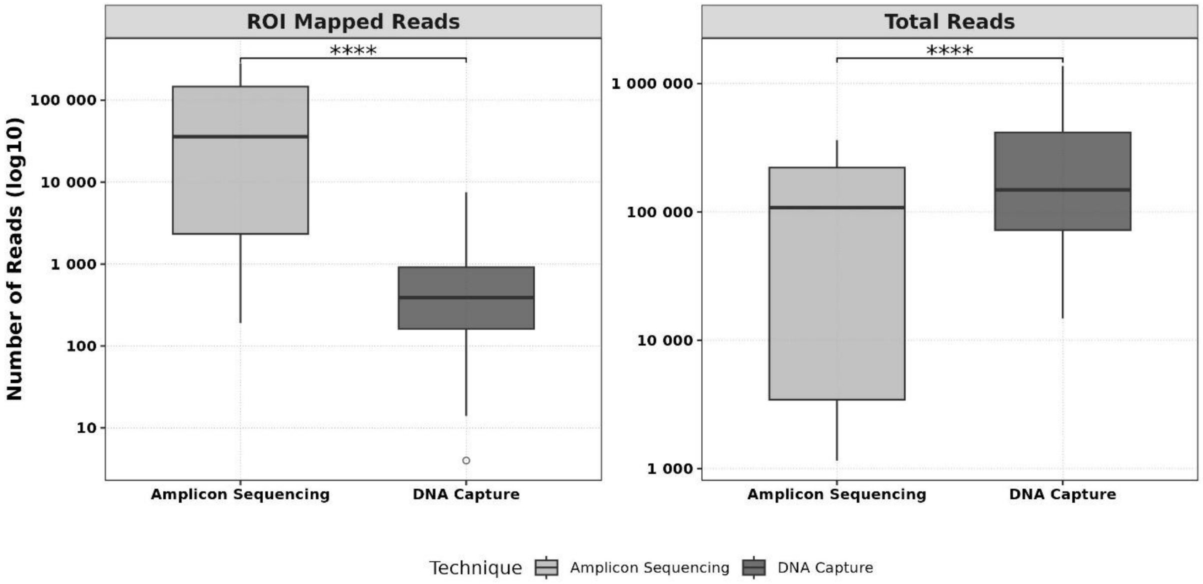


Supplementary figure 1. Comparison of read count produced with the two enrichment approaches (amplicon sequencing and DNA capture).

Total read counts (right) and reads mapped to regions of interest (ROI) (left), the AMR-determining regions were specifically considered. Statistical significance markers are defined as: ****$p<0.0001$.

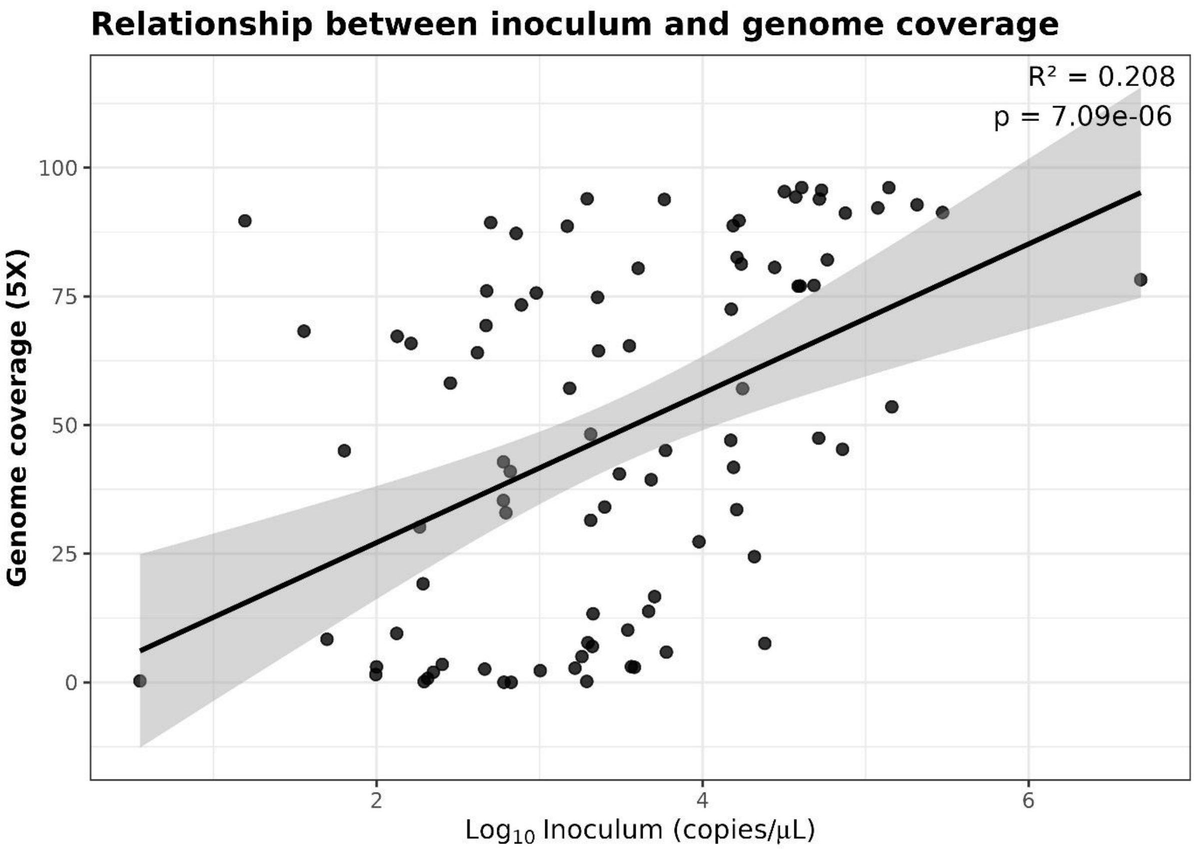


Supplementary figure 2. Relationship between *M. genitalium* load and genome coverage.

The plot indicates a statistically significant positive relationship ($p < 0.001$) as higher inoculum concentrations lead to better breadth of coverage. However, the relationship is relatively weak ($R^2 = 0.208$), suggesting that other additional factors likely play a major role in determining genome coverage.

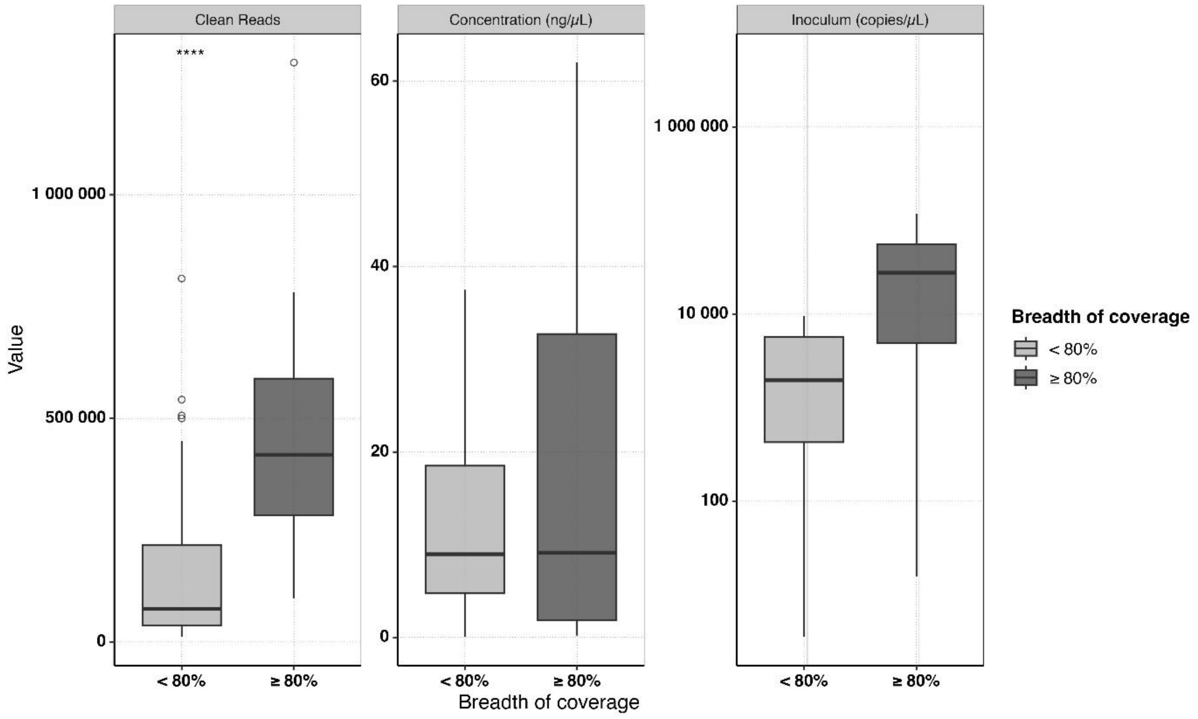


Supplementary figure 3. Impact of clean read count, total DNA concentration, and *M. genitalium* inoculum on *M. genitalium* whole-genome sequencing coverage.

The box plots illustrate the distribution of clean read count, total DNA concentration, and *M. genitalium* inoculum. The data are stratified according to the breath of coverage at 5x (light grey: < 80%; dark grey: ≥ 80%). Statistical significance markers are defined as: ****p<0.0001. The absence of asterisks indicated that no significant differences in distribution were observed between the groups.

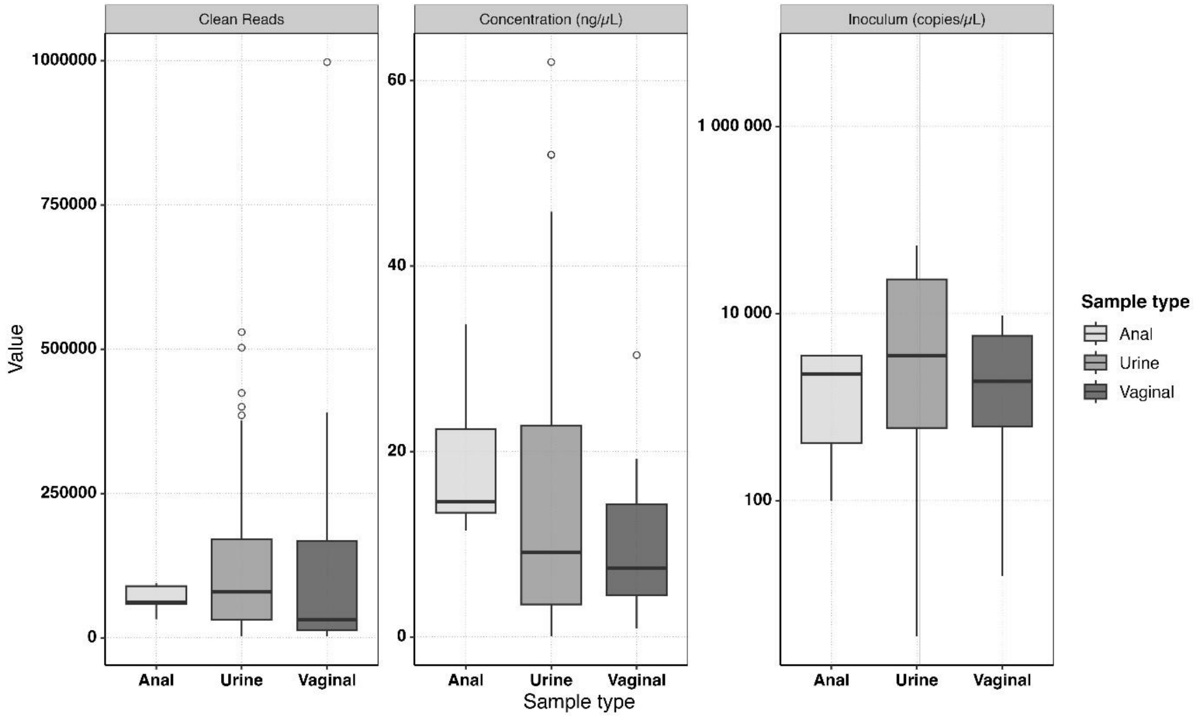


Supplementary figure 4. Clean read count, total DNA concentration, and *M. genitalium* inoculum distribution in specimens across different specimen types.

The horizontal lines within each box represent the median, while the whiskers indicate the range. The absence of asterisks indicated that no significant differences in distribution were observed between the groups.

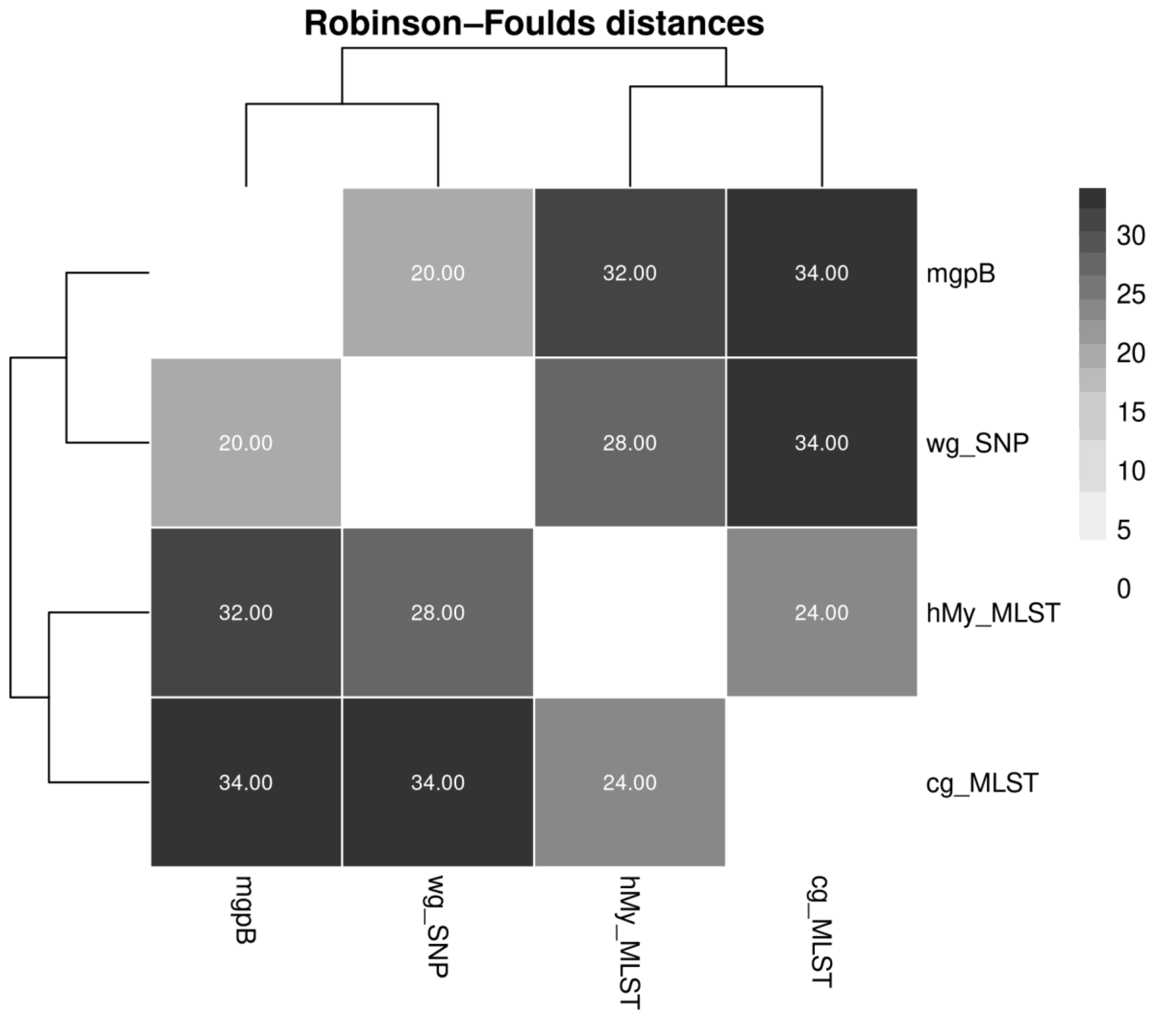


Supplementary Figure 5. Comparison of phylogenetic tree topologies across four typing methods using Robinson-Foulds distances.

The dendrogram represents the hierarchical clustering of the four phylogenetic methods (hMy-MLST, *mgpB*, cg-MLST, and wg-SNP) based on the Robinson-Foulds distance which quantifies topological dissimilarity based on bipartition differences between phylogenetic trees. Methods clustering closer together exhibit greater similarity in their phylogenetic reconstructions. The heatmap visualizes the Robinson-Foulds distance between each pair of methods, with a color gradient ranging from light grey (lower distance value, higher similarity) to dark grey (higher distance value, greater divergence). This combined visualization provides an integrated overview of the relative similarity and divergence among the four methods. The heatmap highlights lower distance values and a relative structural agreement between the two MLST schemes, as well as between the wg-SNP and *mgpB*-based analyses. The heatmap demonstrates the greater divergence of the two MLST schemes from the two other typing methods.

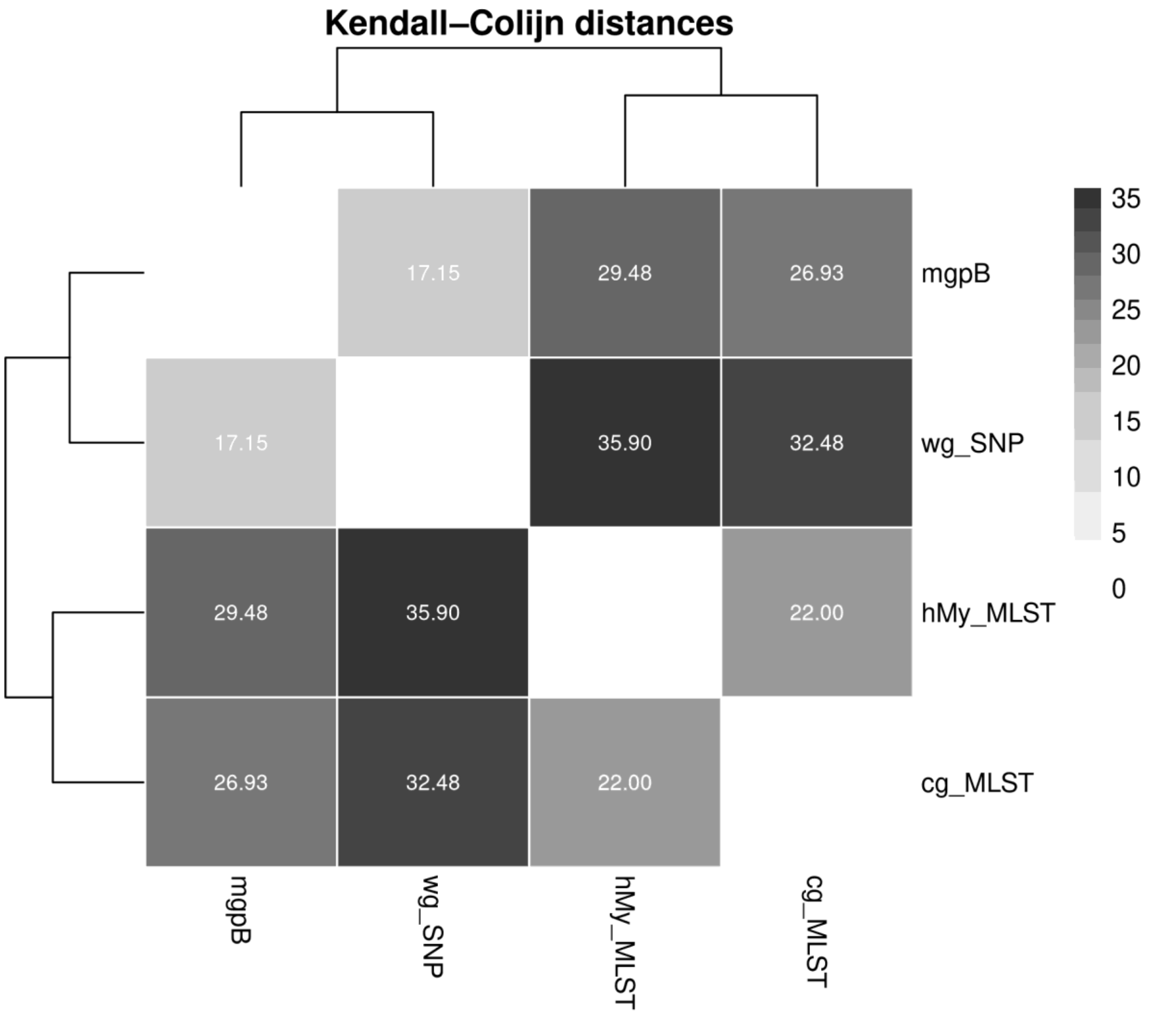


Supplementary Figure 6. Comparison of phylogenetic tree topologies across four typing methods using Kendall-Colijn distances.

The dendrogram represents the hierarchical clustering of the four phylogenetic methods (hMy-MLST, *mgpB*, cg-MLST, and wg-SNP) based on the Kendall-Colijn distance which integrate both tree topology. Methods clustering closer together exhibit greater similarity in their phylogenetic reconstructions. The heatmap visualizes the Kendall-Colijn distance between each pair of methods, with a color gradient ranging from light grey (lower distance value, higher similarity) to dark grey (higher distance value, greater divergence). This combined visualization provides an integrated overview of the relative similarity and divergence among the four methods. The heatmap highlights lower distance values and a relative structural agreement between the two MLST schemes, as well as between the wg-SNP and *mgpB*-based analyses. The heatmap demonstrates the greater divergence of the two MLST schemes from the two other typing methods.
